# Development and validation of prediction models for predicting social care strengths and vulnerability in older people: Cohort study using routine data in Adult Social Care

**DOI:** 10.1101/2025.07.02.25330706

**Authors:** Paul Clarkson, Fiona Lerigo, Glen P. Martin, Leo Wall, Sue Davies, Goran Nenadic, Paul Hine, Catherine Robinson

## Abstract

In Adult Social Care, UK local authorities have statutory responsibilities for assessing needs and delivering services to ensure adults’ wellbeing. Administrative data collected during this process may help local authorities’ compliance with these duties. We developed and internally validated predictive models for older people (>- 60 years) receiving social care for whether they remained at home or were admitted to care homes, two years after index assessments, using administrative data from one English City authority. We enquired whether the right data, to predict older people’s vulnerability to adverse outcome (care home) or evidence their strengths (remaining at home), were present in local authority systems, and if accurate datasets for large numbers of older people could be constructed to allow robust modelling. Logistic regression models were created with binary outcome (remaining at home/entering care homes). Sample size calculations determined the maximum number of candidate predictors we could consider for model development. 20,128 older people in the data cohort indicated we could consider a maximum of 46 candidate predictor parameters. In our final analyses we considered 31 candidate predictor parameters, in the areas of: age, sex, ethnicity, deprivation, housing tenure, abilities in activities of daily living, access to carer, Primary Support Reason, diagnosis of dementia. We used all data for model development and internal validation using cross-validation. Models were robust as to assumptions, with no evidence of overfitting and good predictive accuracy. Circumstances predicting strongly that older people would *not* remain at home were that they were: from other (non-white) ethnic groups, privately rented tenants or other (unstable) tenancies, not able to eat or keep their home habitable, with their Primary Support Reason for personal care, mental health, or support with cognition. With public involvement partners, we created a prototype index for local authority use to inform professional decisions and/or local planning.

## Introduction

In the United Kingdom (UK), Adult Social Care safeguards and protects individuals and provides practical support and care to vulnerable people. Responsibility for Adult Social Care in England lies with 152 local authorities, which have a range of statutory responsibilities to ensure the wellbeing of adults who access and receive social care services [1]. Authorities respond to this by assessing needs and delivering services to hopefully meet those needs. Older people form a large part of the recipients of social care services. Most older people want to stay independent for as long as they can, but many older people underreport their difficulties, often accepting societal assumptions that their circumstances cannot be improved [2]. They may not reveal difficulties to professionals who assess them for the care they might need because of uncertainties over what the responses might be [3]. Likewise, local authority professional assessors may under-detect circumstances driving social care needs amongst older people, sometimes due to inadequacy of assessment documentation or their reticence to probe about circumstances, such as, for example, low mood or incontinence [4]. In both these scenarios, crucial pieces of information might help improve the assessment of future risk to aid decision-making. Ways to gather and use this information unobtrusively, alongside or apart from the face-to-face encounter between older person and assessor, may be beneficial in planning and monitoring the care required and in local authorities’ complying with their legislative duties to promote citizens’ wellbeing.

The process of assessment and service provision in Adult Social Care generates a wealth of routine administrative data on older people’s circumstances, their available resources (such as family carer support) and strengths. However, these data are often underutilised in terms of investigating potential or unmet needs, risks of adverse outcomes or to assist in the longer-term planning of care. This is largely because authorities differ in the sophistication of the data that is generated and held and how it could be used in local evaluation, care planning and research.

There is increasing interest, however, in using the data generated from routine Adult Social Care services for secondary purposes in developing prediction models [5] that can use information about an individual and their environment to estimate their risk of adverse outcomes [6]. Such analyses are relatively more common in healthcare than social care and have been undertaken to inform professional decisions [7,8]. These healthcare prediction models often conceive of predictor variables in the context of deficits or frailty. In the context of Adult Social Care, however, such prediction models could be used to help inform a more positive social vulnerability approach [9], where prediction could identify older people’s daily living difficulties or circumstances that can be mitigated by environmental changes as well as through their own strengths.

The aim of this study was to use routine, administrative data accessed and analysed within Manchester City Council, a large English local authority, to develop and validate a predictive model of social care strengths and vulnerability, two years from initial index assessments as part of the Adult Social Care system. We aimed to discover if and how a prediction model for social care outcomes, using routine data, could be developed, and used to identify older people’s needs and potentially judge threats to their wellbeing. The paper also presents some of the lessons learned from undertaking such predictive modelling in social care, a sector where routine administrative data are not generally used for the purposes of research. We aimed to discover if:

- the right data, to potentially predict older people’s vulnerability to adverse outcomes or evidence their strengths in lessening these, were present in local authority systems.
- accurate, individual-level, datasets for large numbers of older people, representative of those receiving social care, could be constructed to allow robust statistical modelling.
- robust predictive models, statistically predicting outcomes and tested for assumptions, missing data and model fit, could be developed and validated.
- findings could be used to enable a prediction algorithm [10,11] to be made available, as an index for social care, that local authorities could use in planning more appropriate care around older people’s circumstances, resources, and strengths.

## Material and methods

This study was funded by the National Institute for Health and Care Research (NIHR), School for Social Care Research in England as a proof of concept [12]. The reporting of this study adheres to best practice recommendations [13] and followed the Transparent Reporting of a multivariable prediction model for Individual Prognosis or Diagnosis (TRIPOD) guidelines [14]. The concept, data collation and analysis plan had the input of staff from Performance, Research and Intelligence, Manchester City Council who helped develop the application for funding, discussed data availability and access with the research team and helped draft the analysis plan. Our public involvement and engagement partners at Made by Mortals CIC [15], designed innovative, inclusive methods for deliberating on the proposed analyses and development of a predictive index and helped in sharing our learning to other local authority Adult Social Care departments.

### Source of Data and Participants

Data for the development of the prediction model was from the Adult Social Care department of Manchester City Council, administrative data owned by the local authority as a public body, not research data. The Performance, Research and Intelligence team of the council harness the data generated from adult service users’ contacts with the social care system, alongside data from other council departments and local National Health Service (NHS), able to be accessed by them under devolved arrangements between health and social care [16]. There are around 70,000 contacts with Adult Social Care per-year in the city, from a population of over 424,992 adult residents. The data contain the social care information on all adults who access services in the Manchester City local authority area, 32 constituency wards comprising the central urban district within the Greater Manchester region, which covers 10 local authorities in Northwest England.

The study was reviewed and approved by an institutional review board (ethics committee) before the study began (Health Research Authority, Social Care REC: Ref: 23/IEC08/0012). Citizens were informed of the use of their data for other, research, purposes via council Privacy Notices, under Article 9(2)(j) of the UK General Data Protection Regulations (GDPR). For the purposes of the research, a bespoke dataset at the individual person (service user) level, was created, completely anonymised at source, by council data officers, using a data key approach. The agreed data items were anonymised by an officer from the Performance, Research and Intelligence department of the council, with no identifying features remaining in the data shared with researchers. System ID numbers were replaced with a new unique ID; dates of birth were changed to ages and postcodes to Lower Super Output Areas (a geographical area made up of between 400 and 1,200 households). An approved Data Protection Impact Assessment (DPIA) from the council and data sharing agreement governed data collation and dataset access by members of the research team. University research staff were granted access to the dataset (from 31/08/2023) in the council’s secure environment for analysis (researchers went to the data, the data did not come to them), complying with established practice in this type of data access for research [17]. Authors had no access to information that could identify individual participants (data subjects) during or after data collection.

We identified a cohort of older people, aged 60 years and above, from the dataset who had initial contact, care assessments [1], or reviews from the council between 1^st^ April 2021 and 31^st^ March 2023, inclusive. Those under Deprivation of Liberty Safeguards [18] were excluded, due to their non-capacity to consent to council privacy notices concerning access to their data. The index date was taken as the data of first contact assessment for all individuals, with their data followed up until they had the outcome of interest or the end of the cohort period, of 31 March 2023, whichever occurred first.

### Outcomes

The primary outcome was whether older people remained at home (i.e., they were not permanently admitted to a care home) at the end of the cohort period, i.e. as of 31 March 2023. Remaining at home and not entering permanent residential or nursing home care is a key marker of success for social care at home [19] and thus was used as an indicator of older people’s strengths in mitigating any threats to their wellbeing. We also re-ran this model with this outcome reversed, investigating predictors of permanent placement in a care home, an indicator of vulnerability and threats to wellbeing, through the cohort period. This variable was the obverse of the primary outcome, in that it reflects an adverse outcome, albeit entering a care home can be a positive choice for older people and their families. Of all care home admissions in the council dataset, those where the admission was permanent and not temporary, i.e., for respite after hospital discharge or during COVID-19 aftercare arrangements, were used to indicate this outcome of interest.

### Candidate predictor variables

We selected, *a-priori*, potential candidate predictor variables that we hypothesised would be linked to adverse outcomes, like care home admission, or to be indicative of potential threats to older people’s wellbeing, and from the literature [9, 20–23]. These 27 candidate data items (86 predictor parameters) were ones for which we first aimed to investigate whether variables were available in the local authority systems. They are summarised in Box 1. We defined these variables according to data recorded at, or prior to the index date of each individual (i.e., those variables that are available for the first assessments individuals have with the local authority Adult Social Care department). We determined which of these variables were available and could be extracted into a database for analysis, along with reasons for non-availability, through discussions with council data analysts. Additionally, we collapsed categories of some variables in cases of low levels of a categorical variable, and to permit more parsimonious model estimation. Our choice of variables to take forward into the final model analyses were also governed by whether they would be available, or relevant, at the point the prediction model would be applied in practice. For example, service use during the cohort period is a response to circumstances identified at index assessment (in other words, an outcome), so is not relevant here in predicting future needs. In Box 1 we also identify the items that were taken forward as candidates for final model development/analysis, after we had completed this process of combining categories and removing variables irrelevant to prediction.

#### Box 1. Candidate predictor variables evaluated during model development.

##### Demographic

Age, from date of birth, **^b^** in years (continuous), range 60<121 [**^a^**]

Sex * in four categories (Male, Female, Other, Unknown) [**^a^**]

Ethnicity * in 21 categories (Asian or Asian British – Indian, Asian or Asian British – Pakistani, Asian or Asian British – Bangladeshi, Asian or Asian British – Chinese, Asian or Asian British - Any other Asian background, Black, Black British, Caribbean or African – Caribbean, Black, Black British, Caribbean or African – African, Black, Black British, Caribbean or African - Any other Black, Black British or Caribbean background, Mixed or multiple ethnic groups - White and Black Caribbean, Mixed or multiple ethnic groups - White and Black African, Mixed or multiple ethnic groups - White and Asian, Mixed or multiple ethnic groups - Any other Mixed or multiple ethnic background, White - English, Welsh, Scottish, Northern Irish or British, White – Irish, White - Gypsy or Irish Traveller, White – Roma, White - Any other White background, Other ethnic group – Arab, Other ethnic group - Any other ethnic group, No data – Refused, No data - Undeclared or Not known) [**^a^**]

Index of Multiple Deprivation (IMD) [24] (continuous) based on Lower Super Output Area, derived from postcode **^b^** [**^a^**]

##### Living Situation

Living Situation in 2 categories (Alone, with Carer/other family)

Accommodation/Living Situation * in 20 categories (Owner occupier or shared ownership scheme, Tenant, Tenant - private landlord, Settled mainstream housing with family / friends, Supported accommodation / supported lodgings / supported group home, Shared Lives scheme, Approved premises for offenders, Sheltered housing / extra care housing / other sheltered housing, Mobile accommodation for Gypsy / Roma and Traveller communities, Rough sleeper / squatting, Night shelter / emergency hostel / direct access hostel, Refuge, Placed in temporary accommodation by the council (inc. homelessness resettlement), Staying with family / friends as a short-term guest, Acute / long-term healthcare residential facility or hospital, Registered care home, Registered nursing home, Prison / Young offenders institution / detention centre, Other temporary accommodation, Unknown) [**^a^**]

Has unpaid carer **^b^** in 3 categories (Yes, No, unknown) [**^a^**]

##### Functioning

Activities of daily living; Eating & Drinking in two categories, whether have difficulties in (yes, no) [**^a^**]

Activities of daily living; Maintaining Personal Hygiene in two categories, whether have difficulties in (yes, no) [**^a^**]

Activities of daily living; Managing Toilet Needs in two categories, whether have difficulties in (yes, no) [**^a^**]

Activities of daily living; Being Appropriately Clothed in two categories, whether have difficulties in (yes, no) [**^a^**]

Activities of daily living; Maintaining a Habitable Home Environment in two categories, whether have difficulties in (yes, no) [**^a^**]

Primary Support Reason, support reasons in Care Act National Eligibility Criteria **^b^** in 13 categories (Physical Support: Access & mobility only, Physical Support: Personal care support, Sensory Support: Support for visual impairment, Sensory Support: Support for hearing impairment, Sensory Support: Support for dual impairment, Support with Memory & Cognition, Learning Disability Support, Mental Health Support, Social Support: Substance misuse support, Social Support: Asylum seeker support, Social Support: Support for Social Isolation/Other, Social Support: Support to Carer, Unknown) [**^a^**]

Being safe in and around your home in 2 categories (Yes/No)

Making use of facilities and services in the local community in 2 categories (met/unmet)

##### Health conditions likely to incur risk or threats to wellbeing

Visual Impairment **^b^** in 5 categories (Blind/severely sight impaired, Partial sight/sight impaired, No visual impairment, Visual impairment - severity unknown, Unknown

Hearing Impairment **^b^** in 6 categories (Deaf with speech, Deaf without speech, Hard of hearing, No hearing impairment, Hearing impairment - severity unknown, Unknown)

Dementia **^b^** in 3 categories (Yes, No, Unknown) [**^a^**]

Falls in 2 categories (Yes, No)

##### Service use, likely indicative of greater need, or to control for previous service use

Home care in hours per week (continuous)

Day care in hours per week (continuous)

Delivered meals in units per week (continuous)

Delivered equipment/aids in 2 categories (Yes, No)

Transport in units per week (continuous)

Professional support, occupational therapy, social work in units per week (continuous)

Hospital admissions, number within period (continuous)

Accident & Emergency attendances, number within period (continuous)

### Sample Size

As the dataset created was a fixed sample size, calculations were used to determine the maximum number of candidate predictors (prediction parameters) that could be considered for model development (i.e., prior to any data driven predictor selection) [26]. These calculations were performed in the R package pmsampsize [27]. For inputs into these calculations, we used a conservatively proposed c-statistic of 0.7 (i.e., level of discrimination predictive performance that we expected the models to reach), a target shrinkage of 0.9, and an outcome prevalence of 0.96 (remaining at home) and 0.04 (those in care homes) taken from our development data.

For our fixed available sample size (20,128) the sample size calculations indicated we could consider a maximum of 46 candidate predictor parameters (which results in a minimum required sample size of 19,770 and 20,064 for the primary and secondary outcome, respectively). In our final analyses (Tables 1 and 2) we eventually considered 31 candidate predictor parameters.

**Table 1.** Demographic and other characteristics of cohort (n=20,218) for model development and validation. Values are numbers (percentages) unless stated otherwise.

| Characteristic | Full cohort |  |
| --- | --- | --- |
| Mean (SD) age | 76.95 (10.23) |  |
| **Sex** |  |  |
| Female | 11683 (58) |  |
| Male | 8514 (42) |  |
| Unknown | 21 (10) |  |
| **Ethnicity** |  |  |
| White British/Irish | 12602 (62) |  |
| Mixed | 142 (0.7) |  |
| Asian or Asian British | 1362 (7) |  |
| Black or Black British | 1067 (5) |  |
| Other Ethnic Groups | 294 (1.5) |  |
| Refused | 496 (2.5) |  |
| Mean (SD) Index of Multiple Deprivation, min-max | 43.67 (16.35) | 8.15-77.77 |
| **Accommodation Tenure (n=12,373)** |  |  |
| Owner occupier | 4750 (38) |  |
| Private tenant | 539 (4) |  |
| Housing Association tenant | 5006 (40) |  |
| Council tenant | 823 (7) |  |
| Sheltered Housing | 356 (3) |  |
| Residential Care | 530 (4) |  |
| Other | 367 (3) |  |
| **Activities of daily living difficulties Eating (n=4,856)** |  |  |
| Yes | 3794 (78) |  |
| No | 1062 (22) |  |
| **Activities of daily living difficulties Personal Hygiene (n=4,882)** |  |  |
| Yes | 3978 (81) |  |
| No | 904 (19) |  |
| **Activities of daily living difficulties Managing Toilet Needs (n=4,825)** |  |  |
| Yes | 2728 (57) |  |
| No | 2097 (43) |  |
| **Activities of daily living difficulties Appropriately Clothed (n=4,834)** |  |  |
| Yes | 3399 (70) |  |
| No | 1435 (30) |  |
| **Activities of daily living difficulties Maintaining Home Environment (n=4,833)** |  |  |
| Yes | 3597 (74) |  |
| No | 1236 (26) |  |
| **Has unpaid carer (n=20,218)** |  |  |
| Yes | 14375 (71) |  |
| No | 5843 (29) |  |
| **Primary Support Reason (n=7,350)** |  |  |
| Physical Support - Access and Mobility Only | 2251 (31) |  |
| Physical Support - Personal Care Support | 2963 (40) |  |
| Mental Health Support | 316 (4) |  |
| Support with Memory and Cognition | 213 (3) |  |
| Sensory Support - Support for Visual Impairment | 176 (2) |  |
| Sensory Support - Support for Hearing Impairment | 359 (5) |  |
| Sensory Support - Support for Dual Impairment | 140 (2) |  |
| Learning Disability Support | 312 (4) |  |
| Social Support - Support for Social Isolation / Other | 49 (1) |  |
| Social Support - Substance Misuse Support | 104 (1.4) |  |
| Social Support - Asylum Seeker Support | <10<sup>a</sup> |  |
| Social Support - Support to Carer | 466 (6) |  |
| **Dementia (n=20,218)** |  |  |
| Yes | 1807 (9) |  |
| No | 18411 (91) |  |
| **Remains at home (n=20,218)** |  |  |
| Yes | 19505 (96) |  |
| No (care home admission) | 713 (4) |  |

<sup>a</sup> Supressed due to low numbers

### Missing Data

The percentage of missing values across the variable categories included in analysis varied between 0 and 76%. Table S1 in the Supplemental Materials gives the missing data rates of each variable. Many older people had no scores for some variables, as unless the local authority progressed their initial contact to a full Care Act [1] assessment, these variables were not measured. Data was inspected using graphical plots generated by the VIM 6.2.2 package [28], no pattern of missing data differentiated any socio-demographic group of older people. We used multiple imputation to create and analyse five multiple imputed datasets. Methodologists currently regard multiple imputation as the preferred technique for management of missing data because it improves accuracy and statistical power relative to alternative missing data techniques. Graphical checks were performed on continuous variables to observe the relationship between each predictor and outcome variable. Incomplete variables were imputed under fully conditional specification, using the default settings of the mice 3.0 package [29]. Fully conditional specification was used, as in the mice package an imputation method is chosen dependent on the measurement level of each variable and a fully conditional specification is considered suitable where linear relationships are observed between each continuous predictor variable and the outcome variable. Visual plots were inspected to ensure each variable had achieved acceptable imputation. The parameters of substantive interest were estimated in each imputed dataset separately and combined using Rubin’s rules [30].

### Statistical Analysis Methods

As recommended [26], we used all the data for model development and internal validation using cross-validation. This is an iterative process where the full dataset was split into five random folds of equal size. In each step, a fold was left out with the models developed on the remaining folds that was then validated on the excluded fold; this process then repeats at each stage.

For model development, two models were developed. First, we fitted a logistic regression model to the binary outcome of whether older people remained at home versus not (i.e., admitted permanently to care home or other care facility) within two years of the first index contact. Second, we fitted a logistic regression model to this outcome (reversed), treated as a binary outcome of placed in a care home (vs. not placed in care home) after first index contact assessment. For all modelling, we considered all covariates simultaneously as main effects and model assumptions were tested post-estimation.

For validation of each model (in each cycle of the cross-validation process), predictive performance was assessed using metrics of calibration (agreement between observed and expected outcomes) and discrimination (ability of the models to differentiate those who will experience the outcomes from those who will not). We provided calibration plots, assessments of calibration-in-the-large and calibration slope, and c-statistics for discrimination [31]. All performance results were summarised across the cycles of the cross-validation processes, including calculation of 95% confidence intervals. For graphical summaries of predictive performance (calibration plots), we report the calibration plots for each of five iterations in the cross-validation analyses separately. Calibration plots show the observed outcome proportion against the expected outcome proportion from the model, with a perfectly calibrated model having a calibration plot along the y=x line.

All analyses were performed using R version 4.3.1 “Beagle Scouts” [32], along with the packages: broom [33], car [34], dplyr [35], forcats [36], mice [37], predRupdate [38], readxl [39], tidyverse [40], VIM [41], and writexl [42].

## Results

### Overall study cohort

There were 20,128 older people overall in the study cohort who had received at least a contact assessment from the local authority with many (75%) going on to receive subsequent care needs assessments and reviews.

### Data availability

From the list of candidate predictor variables (Box 1) the following functioning variables were unavailable: ‘being safe in and around home’ and ‘making use of facilities and services in the local community’. These were items that would have been drawn from the council strengths and needs assessment, but which were not mandatorily populated in the routine administrative data. In terms of health conditions, falls were not available, being sometimes recorded in primary care, but not mandatorily as routine data available to the council. For service use, meals, transport, and professional support were not available, being not recorded in council systems for contracted agencies (meals, transport) or not systematically available in routine data (professional support).

### Baseline characteristics

Table 1 shows the baseline characteristics of the cohort. The mean age of older service users was 77 years. The majority (58%) were female and White British/Irish (62%). Manchester City residents were living in areas of high deprivation (mean IMD of 44) with 40 per cent living as housing association tenants, and 38 per cent as owner occupiers. In terms of functioning at index assessment, there were significant difficulties with Activities of Daily Living experienced by older users in the domains of eating (78% had difficulties, unable to function without help), personal hygiene (81%), managing the toilet (57%), being appropriately clothed (70%), and maintaining the home (74%). The majority (71%) had an unpaid family carer they could call on for support. In terms of the Primary Support Reason they approached the social care authority for, mostly this was for personal care (40%) and access and mobility (31%). Around 9 per cent of older users had a diagnosis of dementia.

### Predictor variables

The variables eventually included in the models were therefore ones for which information was available and relevant to the concept of identifying risks of future harms (or strengths) in a prediction model. Therefore, those unavailable, above, were excluded from the models. Also, all service use variables (Box 1) were excluded from the modelling, as they were not relevant for predicting future outcomes, as individuals’ service use is itself an (intermediate) outcome. Tables 2 and 3 outline all eventual predictor variables and their candidate predictor parameters included in the final model analyses.

**Table 2.** Prediction model of ‘remaining at home’ (as opposed to entering care homes).

| **Term** | **Estimate (log odds)** | **St. Error** | **95% CI low** | **95% CI high** | **p. value** |
| --- | --- | --- | --- | --- | --- |
| (Intercept) | 4.188581 | 0.585092 | 3.041801 | 5.335360 | 0.000000 |
| Age | 0.006188 | 0.006015 | -0.005600 | 0.017976 | 0.306659 |
| Sex Female | 0.046401 | 0.099936 | -0.149474 | 0.242276 | 0.642589 |
| Ethnicity Mixed | 0.022868 | 0.484477 | -0.926707 | 0.972443 | 0.962383 |
| Ethnicity Asian or Asian British | 1.138768 | 0.284341 | 0.5814605 | 1.696075 | 0.000081 |
| Ethnicity Black or Black British | 0.721807 | 0.237556 | 0.256198 | 1.187416 | 0.002522 |
| Ethnicity Other | -0.35088 | 0.326862 | -0.991524 | 0.289773 | 0.285161 |
| Ethnicity Missing | -0.19034 | 0.314277 | -0.806318 | 0.425648 | 0.546090 |
| Index of Multiple Deprivation (IMD) | 0.008416 | 0.003819 | 0.000930 | 0.015901 | 0.035288 |
| Tenure Private tenant | -0.09096 | 0.359679 | -0.795932 | 0.614010 | 0.801336 |
| Tenure Housing Association tenant | -0.1256 | 0.199759 | -0.517125 | 0.265929 | 0.538641 |
| Tenure Council tenant | 0.360261 | 0.405354 | -0.434231 | 1.154754 | 0.382786 |
| Tenure Sheltered Housing | -0.54095 | 0.397759 | -1.320557 | 0.238655 | 0.197312 |
| Tenure Residential Care | -2.95531 | 0.164514 | -3.277758 | -2.632862 | 0.000000 |
| Tenure Other | -1.47383 | 0.247624 | -1.959170 | -0.988484 | 0.000000 |
| ADL Not able to Eat without difficulty | -0.2086 | 0.240774 | -0.680513 | 0.263321 | 0.399419 |
| ADL Not able to Personal Hygiene without difficulty | 0.114412 | 0.240772 | -0.357501 | 0.586325 | 0.636247 |
| ADL Not able to Toilet without difficulty | -0.22915 | 0.185072 | -0.591888 | 0.133595 | 0.237274 |
| ADL Not able to Dress without difficulty | -0.43826 | 0.225733 | -0.880693 | 0.004180 | 0.066617<sup>a</sup> |
| ADL Not able to Keep Home Habitable without difficulty | -0.42642 | 0.24324 | -0.903168 | 0.050331 | 0.108184 |
| No Access to a carer | -0.25931 | 0.109065 | -0.473073 | -0.045537 | 0.017509 |
| Primary Support Reason Physical Support | -0.36039 | 0.128976 | -0.613180 | -0.107594 | 0.005319 |
| Primary Support Reason Mental Health Support | -1.43507 | 0.213817 | -1.854150 | -1.015988 | 0.000000 |
| Primary Support Reason Memory and Cognition | -0.35633 | 0.286978 | -0.918807 | 0.206144 | 0.219210 |
| Primary Support Reason Visual Impairment | 0.354602 | 0.480941 | -0.588042 | 1.297247 | 0.462685 |
| Primary Support Reason Hearing Impairment | 0.01328 | 0.304823 | -0.584173 | 0.610733 | 0.965273 |
| Primary Support Reason Dual Impairment | -0.36689 | 0.445779 | -1.240612 | 0.506840 | 0.414019 |
| Primary Support Reason Learning Disability | 0.543922 | 0.286407 | -0.017436 | 1.105279 | 0.059742<sup>a</sup> |
| Primary Support Reason Social Isolation | 0.527212 | 1.162209 | -1.750718 | 2.805142 | 0.653678 |
| Primary Support Reason Substance Misuse | 0.007838 | 0.53292 | -1.036686 | 1.052361 | 0.988281 |
| Primary Support Reason Support to Carer | 2.663495 | 1.031149 | 0.642442 | 4.684547 | 0.009860 |
| Cognitive Impairment/dementia (diagnosis) yes | -0.19968 | 0.131292 | -0.457016 | 0.057647 | 0.128429 |

N=20,128 older people (60 years and over) receiving Adult Social Care support over two years; index request for service or contact during April-May 2021, followed up for 24 months, until March 2023. Outcome events: remaining at home N= 19505 (96%), resident in care homes N= 713 (4%)
Yellow rows are significant at $p < 0.05$ ; <sup>a</sup>Nears significance.
ADL=Activities of Daily Living.
The model is a glm generalised linear model (binary logistic). The assumptions for a logistic regression were checked for: appropriate outcome i.e., binary, independence of variables, large enough sample size, linearity of predictors and log odds, and multicollinearity (with gVIF - generalised VIF to ensure no variable introduced a VIF above 5).

### Model evaluation

Tables 2 and 3 show the coefficients of our final models for remaining at home or entering care homes respectively. For the model of remaining at home (Table 2) the following were significant predictors: Ethnicity Asian or Asian British, Black or Black British (more likely to remain at home); Living in area of high deprivation (more likely to remain at home); living in residential care or ‘other’ tenancy – including unstable accommodation (less likely to remain at home); No access to carer (less likely to remain at home)’ Primary Support Reason of needing physical or mental health support (less likely to remain at home) or support to carer (more likely to remain at home). For the model of entering care homes (Table 3) the following were significant predictors: Ethnicity Black or Black British (less likely to enter care homes) or other ethnicity (more likely to enter care homes); living in private tenancy (more likely to enter care homes); living in residential care or other tenancy (more likely to enter care homes); Activities of Daily Living (ADL) unable to eat and keep the home habitable without difficulty (more likely to enter care homes); Primary Support Reason being need for physical, mental health support or support with memory and cognition (more likely to enter care homes).

**Table 3.** Prediction model of risk of care home admission (not ‘remaining at home’).

| **Term** | **Estimate (log odds)** | **St. Error** | **95% CI low** | **95% CI high** | **p. value** |
| --- | --- | --- | --- | --- | --- |
| (Intercept) | -4.557854 | 0.546177 | -5.62836 | -3.48734 | 0.000000 |
| Age | -0.006215 | 0.005478 | -0.01695 | 0.004521 | 0.243958 |
| Sex Female | 0.024653 | 0.103842 | -0.17887732 | 0.22818 | 0.812572 |
| Ethnicity Mixed | 0.071110 | 0.103842 | -0.13242032 | 0.274640 | 0.812572 |
| Ethnicity Asian or Asian British | -1.266794 | 0.456061 | -2.16067356 | -0.372914 | 0.876992 |
| Ethnicity Black or Black British | -0.602220 | 0.31722 | -1.2239712 | 0.019531 | 0.000087 |
| Ethnicity Other | 0.169991 | 0.243006 | -0.30630076 | 0.646282 | 0.013949 |
| Ethnicity Missing | -0.142931 | 0.349327 | -0.82761192 | 0.541749 | 0.636800 |
| Index of Multiple Deprivation (IMD) | -0.009145 | 0.327451 | -0.65094896 | 0.632658 | 0.679883 |
| Tenure Private tenant | 0.2393726 | 0.003233 | 0.23303592 | 0.245709 | 0.005209 |
| Tenure Housing Association tenant | 0.1624988 | 0.39566 | -0.6129948 | 0.937992 | 0.575564 |
| Tenure Council tenant | -0.073557 | 0.157711 | -0.38267056 | 0.235556 | 0.301278 |
| Tenure Sheltered Housing | 0.8054793 | 0.296915 | 0.2235259 | 1.387432 | 0.777530 |
| Tenure Residential Care | 2.865961 | 0.247525 | 2.380812 | 3.35111 | 0.001215 |
| Tenure Other | 1.432942 | 0.170124 | 1.09949896 | 1.766385 | 0.000000 |
| ADL Not able to Eat without difficulty | 0.538441 | 0.290271 | -0.03049016 | 1.107372 | 0.000106 |
| ADL Not able to Personal Hygiene without difficulty | -0.385531 | 0.263049 | -0.90110704 | 0.130045 | 0.059900<sup>a</sup> |
| ADL Not able to Toilet without difficulty | 0.277418 | 0.297527 | -0.30573492 | 0.860570 | 0.219425 |
| ADL Not able to Dress without difficulty | 0.732575 | 0.193456 | 0.35340124 | 1.111748 | 0.182303 |
| ADL Not able to Keep Home Habitable without difficulty | 0.368280 | 0.267186 | -0.15540456 | 0.891964 | 0.015232 |
| No Access to a carer | 0.257110 | 0.24025 | -0.21378 | 0.728 | 0.144941 |
| Primary Support Reason Physical Support | 0.422323 | 0.109081 | 0.20852424 | 0.636121 | 0.019837 |
| Primary Support Reason Mental Health Support | 1.628625 | 0.163591 | 1.30798664 | 1.949263 | 0.015068 |
| Primary Support Reason Memory and Cognition | 0.472469 | 0.209639 | 0.06157656 | 0.883361 | 0.000000 |
| Primary Support Reason Visual Impairment | 0.319450 | 0.27244 | -0.2145324 | 0.853432 | 0.083710 |
| Primary Support Reason Hearing Impairment | 0.486897 | 0.383585 | -0.2649296 | 1.238723 | 0.407499 |
| Primary Support Reason Dual Impairment | 0.144464 | 0.30265 | -0.44873 | 0.737658 | 0.105214 |
| Primary Support Reason Learning Disability | -0.346596 | 0.439455 | -1.2079278 | 0.514735 | 0.764383 |
| Primary Support Reason Social Isolation | -0.433345 | 0.391162 | -1.20002252 | 0.333332 | 0.392372 |
| Primary Support Reason Substance Misuse | 0.574834 | 1.089408 | -1.56040568 | 2.710073 | 0.643513 |
| Primary Support Reason Support to Carer | -14.70269 | 0.533333 | -15.7480227 | -13.657357 | 0.305576 |
| Cognitive Impairment/dementia (diagnosis) yes | 0.212391 | 285.9956 | -560.338985 | 560.763767 | 0.958981 |

N=20,128 older people (over 60 years) receiving Adult Social Care support over two years; index request for service or contact during April-May 2021, followed up for 24 months, until March 2023. Outcome events: resident in care homes N= 713 (4%), remaining at home N= 19505 (96%)
Yellow rows are significant at $p < 0.05$ ; \*Nears significance.
ADL=Activities of Daily Living.
The model is a glm generalised linear model (binary logistic). The assumptions for a logistic regression were checked for: appropriate outcome i.e., binary, independence of variables, large enough sample size, linearity of predictors and log odds, and multicollinearity (with gVIF - generalised Variance Inflation Factor to ensure no variable introduced a VIF above 5).

Both models were robust as to assumptions for logistic regression, i.e. binary, independence of variables, large enough sample size, linearity of predictors and log odds, and multicollinearity.

### Model validation

Tables 4 and 5 present the predictive performance results from the internal cross-validation process for each model. It can be seen, in the data reflecting the full models, that calibration-in-the-large/pooled c-statistic (calibration intercept = 0) is good and no evidence of overfitting (calibration slope inside range 0.9-1.1). Low Brier scores indicated good predictive accuracy.

**Table 4.** Model of ‘remaining at home’: Validation – calibration indices. ^a^.

|  | Estimate | SE | CI 95% lower | CI 95% upper |
| --- | --- | --- | --- | --- |
| Pooled c-statistic | 0.8386 | 0.0184 | 0.8025 | 0.8747 |
| Pooled intercept | **0.0166** | 0.09544 | -0.17046 | 0.2037 |
| Pooled slope | **1.0120** | 0.06012 | 0.894167 | 1.12983 |
| Pooled Cox Snell | 0.0706 |  |  |  |
| Pooled Nagelkerke | 0.2768 |  |  |  |
| Pooled Brier Score | 0.0272 |  |  |  |

<sup>a</sup> The figures above are based on the pooled estimates of 5 imputations. The pooled estimates have been calculated using Rubin’s Rules (RR). It can be seen (bold), in the data that reflects the full regression model, that calibration-in-the-large (calibration intercept = 0) is good and no evidence of overfitting (calibration slope inside range 0.9-1.1).

**Table 5.**
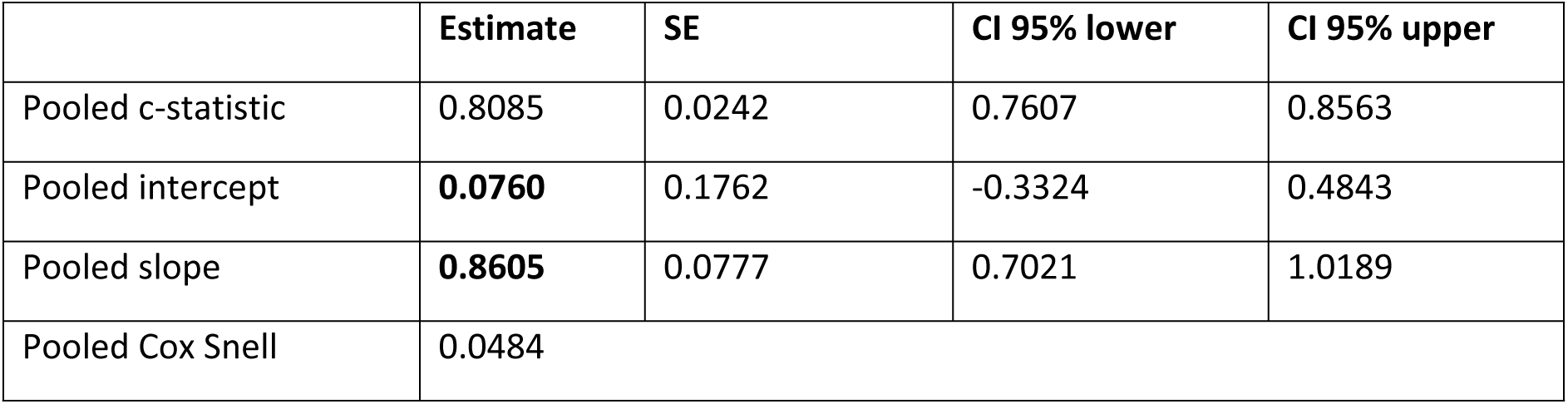
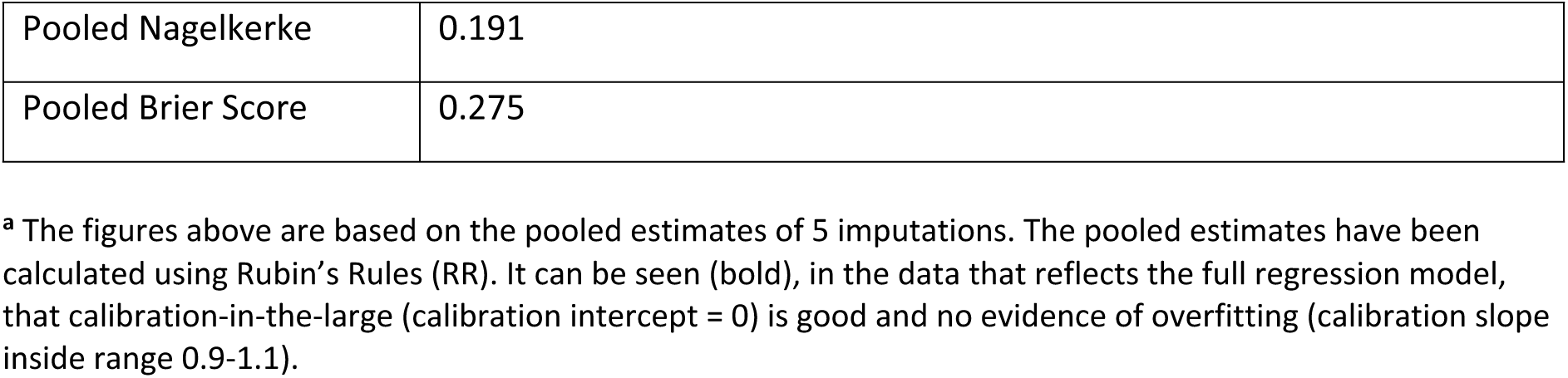
Model of entering care home: Validation – calibration indices. ^a^.

|  | Estimate | SE | CI 95% lower | CI 95% upper |
| --- | --- | --- | --- | --- |
| Pooled c-statistic | 0.8085 | 0.0242 | 0.7607 | 0.8563 |
| Pooled intercept | **0.0760** | 0.1762 | -0.3324 | 0.4843 |
| Pooled slope | **0.8605** | 0.0777 | 0.7021 | 1.0189 |
| Pooled Cox Snell | 0.0484 |  |  |  |

|  |  |
| --- | --- |
| Pooled Nagelkerke | 0.191 |
| Pooled Brier Score | 0.275 |

<sup>a</sup> The figures above are based on the pooled estimates of 5 imputations. The pooled estimates have been calculated using Rubin's Rules (RR). It can be seen (bold), in the data that reflects the full regression model, that calibration-in-the-large (calibration intercept = 0) is good and no evidence of overfitting (calibration slope inside range 0.9-1.1).

Fig S1. in supplementary information, presents calibration plots of observed vs predicted probability for the models of remaining at home in each of the five cycles of the cross-validation process. These show that the models were well calibrated across the full risk range.

### Developing a predictive index

Results from these analyses were shared with the council and in an interactive, immersive workshop event run by our public involvement and engagement partners [43]. An audience of stakeholders (council data intelligence officers, service managers, social workers, charity members, researchers, and older people drawing on social care) deliberated on the form a social care predictive index could take. It was envisaged that this could be to aid professional judgement in individual cases and/or support service planning or forecasting at a population level. It could also put the older person in control of their care, in conversations with professionals, by using the data from the index as a feedback mechanism to guide joint decision-making. An index was thought to be broadly acceptable to stakeholders, but with some caveats. Basing an index on the right data, with comprehensive analysis and assurances of data security were seen as important.

We developed the index with the feedback from the workshop event in mind. From our analyses, we determined the contributions of each predictor variable to the outcome from the model of risk of care home admission. We then used these to weight individual predictions, (by odds ratio estimates), for older people with different characteristics, in an exemplar interactive index. We used R and a Shiny package to create this [44]. Our exemplar index, predicting individuals’ risk of no longer remaining at home, is openly accessible (at https://scs-10.shinyapps.io/Visable/), giving an output of calculated risk, based on overall odds ratio, interpreted as the degree of risk of being in a care home if the person has the circumstances considered, as opposed to not having them. An odds ratio greater than one indicates increased risk, while a value less than one indicates decreased risk (protective effect). From this index, Figs S2 and S3 in the Supplemental Materials show two examples, highlighting the risk of care home admission, for relatively high and low risk individuals respectively.

Supplementary Fig S2 shows a 70-year woman from an inner-city area [45], in a private tenancy, with ‘other’ (non-white) ethnicity. In terms of need, they were not able to feed themselves or look after their home, but were able to bathe and groom, use the toilet and dress themselves. They did not have access to a carer and their Primary Support Reason on access to social care was for ‘mental health support’. They were known to have a dementia diagnosis. Their overall risk (odds ratio) is 2.5; they are 150% more likely to be in a care home two years after initial assessment than someone without these characteristics.

Supplementary Fig S3 shows a 61-year-old white man, an owner-occupier from a well-resourced outer-city area. They were able to undertake all daily living activities but had difficulties looking after their home. They had access to a carer and their Primary Support Reason was for access and mobility. They did not have dementia. Their overall risk (odds ratio) is 0.26; they are 26% less likely to be at risk of being in a care home.

It is not recommended that this prototype index yet be used to support operational decision making in individual cases in local authority Adult Social Care departments. The analyses presented here require further external validation in other local authorities before this is appropriate. Work is planned on an external validation in other local authorities in the UK, the results of which will be presented separately. Work is also underway to investigate how the index could potentially be reconfigured to assist service planning at a population level, rather than individual risk profiling.

## Discussion

We developed and validated statistical models from routine administrative Adult Social Care data in a local authority in Greater Manchester, an urban, city authority with a diverse population of older people requiring access to social care support. From these, we provided robust, well-calibrated models for predicting which sorts of older people’s circumstances predicted the likelihood of them either remaining at home or entering care homes two-years after initial assessment. We found circumstances predicting strongly that older people would *not* remain at home, and enter care homes, were: that they belonged to other (non-white) ethnic groups; that they were privately rented tenants or living in other (unstable) tenancies; that, in terms of their activities of daily living, they were not able to eat or keep their home habitable without difficulty; that their Primary Support Reason was for personal care, mental health, or support with memory and cognition.

These findings have broad face validity according to variables from research investigations into the factors most likely to predict the living circumstances of older people from initial assessments of their needs [20–23]. Particularly not having access to a carer (showing a significant negative relationship in our remaining at home model) and requiring personal care, mental health support and support with memory and cognition would be expected to be important in determining older people’s risks of entering care homes. Also, very important at initial assessment is the level of performance in the activities of daily living, whose relationship with older people’s later living situation is evidenced in previous research enquiry. Abilities in activities of daily living follow a set pattern through life, those reflecting basic functions essential to survival – such as feeding – are acquired first with more complex learned activities – such as dressing – acquired last [46]. In aging or as long-term conditions are acquired, this process is reversed; the more basic functions are retained the longest and so cases where older people experience difficulties with these should be signalled for special attention [47]. Our finding of activity difficulties with eating being a significant predictor of the likelihood of care home admission reinforces the use of such a data item as integral to assessing risk in social care.

However, although these findings appear more-or-less established from research studies, one advantage of our study is that the administrative data are from large numbers of real, routine cases in Adult Social Care departments, that have complied with eligibility criteria for support and exhibit a range of care needs, and from operational data of users’ journeys through the care system. The data are thus representative of the population of older social care users receiving support from the local authority. Our sample size was more than adequate and predictor-outcome relationships, on the whole, are as expected from the primary research evidence. On the other hand, one limitation is our finding from area deprivation (drawn from anonymised post-code linkage), which appeared counter- intuitive, as those in areas of greater deprivation were *more* likely to remain at home. This could be explained by the distribution of the data, particularly with Manchester City having a preponderance of higher deprived small local areas. And, therefore, there is a much greater effect of higher deprivation on the likelihood of remaining at home; in Manchester, 19,505 (96%) older people remained at home two-years after initial assessment, as opposed to 1,236 (4%) entering care homes on a permanent basis. There is therefore a need to investigate the generalisability of our models across regions of the UK, where the prevalence of deprivation may be lower and so its impact on the likelihood of care home admission may be more pronounced. A further limitation is our omission of some relevant well-known predictors of care home admission [22], for which the data were unavailable. Older people’s feelings of safety at home and the existence of falls are relevant predictors of entering care homes but were unavailable to include in our models.

We were able to develop a prototype index from these analyses and prediction estimates that the council and other local authorities, could potentially use to help inform future professional decisions, service configuration and/or local planning. User, practitioner and manager views on such an index were that it was broadly acceptable but that it must be based on the right data, that the data and analysis was trusted, and that the data were secure. We provide examples from this index, which can be used to inform discussion about its future development. As stated, our study was a proof-of-concept to examine whether the right data in this setting could be available and that models could be run on the data that were well calibrated and could reliably predict our chosen outcomes. We would not recommend our prototype decision-making tool yet be used routinely by local authorities to judge risk in individual older people. Further external validation in other local authorities is needed, particularly those with differences, importantly, in deprivation levels, rurality, and ethnic mix that are different to those in Manchester. In addition, we would like to improve the useability of the tool for social care professionals and managers in terms of the information it can provide, for those largely unfamiliar with statistical language and outputs.

Adult social care practice in England does not presently capitalise enough on the wealth of routine data existing and generated within local authority Adult Social Care. Our study showed that much of the data most useful in predicting what might happen to older users in their care trajectories is now currently being collected by local authorities. These data items, including those of Primary Support Reason, carer access, accommodation-type, and ethnicity, are those now mandated in the Client Level Data (CLD) set, and so will be collectable by local authority performance and intelligence teams. The CLD is now planned to be available nationally, for all local authorities, also for researchers to eventually access and use [48, 49]. Therefore, this can be used in future to undertake more sophisticated analyses, on users’ journeys through the care system to inform evaluations of how councils are responding to their Care Act [1] responsibilities. Our analyses point the way to how this could successfully be done, in partnership with local authority Adult Social Care departments.

## Data Availability

Data cannot be shared publicly because it is owned and controlled by Manchester City Council, a Local Authority in England in compliance with their duties under the Care Act 2014 and other UK legislation. It is administrative data and not generally available for research or for other research investigators to use in the UK, or in any other jurisdiction.

https://www.madebymortals.org/for-datas-sake-an-interactive-event-exploring-a-new-social-care-index/

https://doi.org/10.48420/25703187.v2

https://scs-10.shinyapps.io/Visable/

## Acknowledgements

We thank the staff at Performance Research and Intelligence, Manchester City Council for collating the administrative data used in the study.

## Supporting information

**S1 Checklist. TRIPOD checklist: Prediction model development and validation.** (PDF)

**S1 Table. Missingness in variables used in the logistic models.**

**S1 Fig. Calibration plots of observed vs predicted probability for the models of remaining at home.** (PNG)

**S2 Fig. Example risk calculation for relatively high-risk case.** (PDF)

**S3 Fig. Example risk calculation for relatively low risk case.** (PD

## Footnotes

a considered as candidates for final model development/analysis after undertaking process of category-combining and removing irrelevant variables.

b Items included in the Adult Social Care Client Level Data (CLD); the mandatory data set that local authorities began to submit to NHS England from July 2023 [25]

## References

[1] UK Government. Care Act 2014, c.23; Available from: http://www.legislation.gov.uk/ukpga/2014/23/pdfs/ukpga_20140023_en.pdf

[2] Philp I. No time to lose. In: Goodwin J, Rossall P, editors. Improving Later Life. London: Age UK; 2012 pp. 58–61.

[3] Howse K, Ebrahim S, Gooberman-Hill R. Help-avoidance: why older people do not always seek help. Rev Clin Gerontol. 2005; 14: 63–70. 10.1017/S0959259804001212

[4] Clarkson P, Abendstern M, Sutcliffe C, Hughes J, Challis D. Reliability of needs assessments in the community care of older people: Impact of the Single Assessment Process in England. Journal of Public Health. 2009; 31: 521–529. 10.1093/pubmed/fdp035

[5] Riley RD, van der Windt D, Croft P, Moons KGM. Prognosis research in healthcare concepts, methods, and impact. Oxford: Oxford University Press; 2019.

[6] NHS England Transformation Directorate. AI in adult social care: Innovation to improve outcomes and increase choices for adult social care service users. 2021; Available from: https://www.nhsx.nhs.uk/ai-lab/explore-all-resources/understand-ai/ai-adult-social-care/

[7] Sessler DI, Sigl J, Manberg PJ, Kelley S, Schubert A, Chamoun NG. (2010) Broadly applicable risk stratification system for predicting duration of hospitalization and mortality. Anesthesiology. 2010; 113: 1026–1037. doi: 10.1097/ALN.0b013e3181f79a8d.

[8] Zhao J, Guo Z, Liu X, Glaser DL, Elkassabany NM, Liu J. The development and application of a risk stratification index system for outpatient shoulder arthroscopy patient management—a single academic center’s experience. JSES Open Access. 2017; 1–4. doi: 10.1016/j.jses.2017.03.005.

[9] Andrew MK, Mitnitski AB, Rockwood K. Social vulnerability, frailty and mortality in elderly people. PLoS One. 2008; 3(5): e2232. doi:10.1371/journal.pone.0002232.

[10] Clift AK, Coupland CAC, Keogh RH. et al. Living risk prediction algorithm (QCOVID) for risk of hospital admission and mortality from coronavirus 19 in adults: national derivation and validation cohort study. BMJ. 2020; 371:m3731; 10.1136/bmj.m3731

[11] Clegg A, Bates C, Young J. et al. Development and validation of an electronic frailty index using routine primary care electronic health record data. Age Ageing. 2016; 45: 353–360; 10.1093/ageing/afw039

[12] Kush R, Alschuler L, Ruggeri R. et al. (2007) Implementing single source: The STARBRITE proof-of-concept study. J Am Med Inform Assoc. 2007; 14: 662–673. doi: 10.1197/jamia.M2157.

[13] Steyerberg EW. Clinical prediction models. New York: Springer; 2009.

[14] Collins GS, Reitsma JB, Altman DG, Moons KGM. Transparent Reporting of a multivariable prediction model for Individual Prognosis or Diagnosis (TRIPOD): the TRIPOD statement. Ann Intern Med. 2015; Jan 6; 162(1): 55–63; doi: 10.7326/M14-0697.

[15] Made by Mortals: Bringing lived experience to life. 2025; Available from: https://www.madebymortals.org/

[16] Malik B, Wells J, Hughes J, Clarkson P, Keady J, Young A, Challis D. Complex care needs and devolution in Greater Manchester: a pilot study to explore social care innovation in newly integrated service arrangements for older people. Aust Health Rev. 2020; 44(6): 838–846. doi:10.1071/AH19168.

[17] Desai T, Ritchie F, Welpton R. Five Safes: designing data access for research. Bristol Business School Working Papers in Economics. 2016; 1601. Available from: https://www2.uwe.ac.uk/faculties/BBS/Documents/1601.pdf

[18] UK Government. Deprivation of liberty safeguards: resources. 2024; Available from: https://www.gov.uk/government/publications/deprivation-of-liberty-safeguards-forms-and-guidance

[19] Wiles JL, Leibing J, Guberman N, Reeve J, Allen RES. The meaning of “aging in place” to older people. Gerontologist. 2011; 52: 357–366. doi:10.1093/geront/gnr098.

[20] Hildon ZJL, Tan CS, Shiraz F. et al. The theoretical and empirical basis of a BioPsychoSocial (BPS) risk screener for detection of older people’s health related needs, planning of community programs, and targeted care interventions. BMC Geriatr. 2018; 18:49. 10.1186/s12877-018-0739-x

[21] Clarkson P, Venables D, Hughes J. et al. Integrated specialist assessment of older people and predictors of care-home admission. Psychological Medicine. 2006; 36: 1011–1021. doi: 10.1017/S0033291706007434.

[22] Luppa M, Luck T, Weyerer S, König H-H, Brähler E, Riedel-Heller SG. Prediction of institutionalization in the elderly. A systematic review. Age ageing. 2010; 39: 31–38. doi: 10.1093/ageing/afp202.

[23] Bardsley M, Billings J, Dixon J, Georghiou T, Lewis H, Teventon, A. Predicting who will use intensive social care: case finding tools based on linked health and social care data. Age and Ageing. 2011; 40: 265–270.

[24] UK Government. Ministry of Housing, Communities and Local Government. The English Indices of Deprivation 2019. Frequently Asked Questions. 2019; Available from: https://assets.publishing.service.gov.uk/government/uploads/system/uploads/attachment_data/file/853811/IoD2019_FAQ_v4.pdf

[25] NHS Digital. Collection of Client Level Adult Social Care Data (No 3) Directions 2023; https://digital.nhs.uk/about-nhs-digital/corporate-information-and-documents/directions-and-data-provision-notices/secretary-of-state-directions/collection-of-client-level-adult-social-care-data-no-3

[26] Riley RD, Ensor J, Snell KIE, Harrell FE, Martin GP, Reitsma JB, et al. Calculating the sample size required for developing a clinical prediction model. BMJ. 2020; Mar 18: m441. doi: 10.1136/bmj.m441.

[27] Package ‘pmsampsize’, October 14, 2022, Version 1.1.2; The Comprehensive R Archive Network; https://cran.r-project.org/

[28] Kowarik A, Templ M. Imputation with the R Package VIM. Journal of Statistical Software. 2016; 74(7): 1–16. 10.18637/jss.v074.i07

[29] van Buuren S, Groothuis-Oudshoorn K. mice: Multivariate Imputation by Chained Equations in R. Journal of Statistical Software. 2011; 45(3): 1–67. 10.18637/jss.v045.i03

[30] Rubin DB. Multiple imputation for nonresponse in surveys. New York: John Wiley & Sons, Inc; 1987.

[31] Steyerberg EW, Vickers AJ, Cook NR, Gerds T, Gonen M, Obuchowski N, et al. Assessing the performance of prediction models: a framework for traditional and novel measures. Epidemiology. 2010; 21(1): 128–138.

[32] R Core Team (2023). R: A Language and Environment for Statistical Computing. R Foundation for Statistical Computing, Vienna, Austria; https://www.R-project.org/

[33] Robinson D, Hayes A, Couch S. broom: Convert statistical objects into tidy tibbles. R package version 1.0.7. 2024; https://CRAN.R-project.org/package=broom

[34] Fox J, Weisberg S. (2019). An R Companion to Applied Regression. Third edition. Thousand Oaks CA: Sage; https://www.john-fox.ca/Companion/

[35] Wickham H, François R, Henry L, Müller K, Vaughan D. dplyr: A grammar of data Manipulation.. R package version 1.1.4. 2023; https://CRAN.R-project.org/package=dplyr

[36] Wickham H. forcats: Tools for Working with Categorical Variables (Factors). R package version 1.0.0. 2023; https://CRAN.R-project.org/package=forcats

[37] van Buuren S, Groothuis-Oudshoorn K. (2011). mice: Multivariate imputation by chained equations in R. Journal of Statistical Software. 2011; 45(3): 1–67. DOI 10.18637/jss.v045.i03.

[38] Martin G, Jenkins D, Sperrin M. predRupdate: Prediction model validation and updating. R package version 0.2.0. 2024; https://CRAN.R-project.org/package=predRupdate

[39] Wickham H, Bryan J. readxl: Read Excel Files. R package version 1.4.5. 2025; https://CRAN.R-project.org/package=readxl

[40] Wickham H, Averick M, Bryan J, Chang W, McGowan LD, François R, et al. “Welcome to the tidyverse.” Journal of Open-Source Software. 2019; 4(43): 1686. doi:10.21105/joss.01686; https://doi.org/10.21105/joss.01686

[41] Templ M, Kowarik A, Alfons A, Cillia G, Prantner B, Rannetbauer W. VIM: Visualization and imputation of missing values. R package version 6.2.2. 2025; https://cran.r-project.org/web/packages/VIM/index.html

[42] Ooms J, McNamara J. writexl: Export data frames to Excel ‘xlsx’ format. R package version 1.5.4. 2025; https://cran.r-project.org/web/packages/writexl/index.html

[43] Made by Mortals. For Data’s Sake: An interactive event exploring a new social care index. 2025; Available from: https://www.madebymortals.org/for-datas-sake-an-interactive-event-exploring-a-new-social-care-index/

[44] Ji X, Kattan MW. Tutorial: development of an online risk calculator platform. Ann Transl Med 2018; 6(3): 46. doi: 10.21037/atm.2017.11.37.

[45] Manchester City Council. Deprivation: data and intelligence. 2024; Available from: https://www.manchester.gov.uk/info/200088/statistics_and_intelligence/2168/deprivation

[46] Williams RG, Johnston M, Willis LA, Bennett AE. Disability: a model and measurement technique. British Journal of Preventative and Social Medicine. 1976; 30: 71–8. doi: 10.1136/jech.30.2.71.

[47] Clarkson, P. What research tells social workers about their work with older people. In Davies M, editors. Social work with adults. London: Palgrave, Macmillan; 2012. p. 300–313.

[48] Social Care Institute for Excellence (SCIE). How data can support better outcomes for people needing care and support, using Client Level Data (CLD). January 2024; Available from: https://www.scie.org.uk/innovation/showcase-webinars/client-level-data-cld/#:∼:text=Client%2DLevel%20Data%20(CLD),under%20the%20Care%20Act%202014.

[49] Arden&GEM. Adult Social Care Client Level Data (CLD), NHS Digital; Available from: https://www.ardengemcsu.nhs.uk/adult-social-care-client-level-data/

